# Genome-wide transcriptomic analysis of intestinal mucosa in celiac disease patients on a gluten-free diet and post gluten challenge

**DOI:** 10.1101/2020.04.17.20067942

**Authors:** Valeriia Dotsenko, Mikko Oittinen, Juha Taavela, Alina Popp, Markku Peräaho, Synnöve Staff, Jani Sarin, Francisco Leon, Jorma Isola, Markku Mäki, Keijo Viiri

## Abstract

**Background & Aims:** Gluten challenge studies are instrumental in understanding the pathophysiology of celiac disease. Our aims in this study were to reveal early gluten-induced transcriptomic changes in duodenal biopsies and to find tools for clinics.

**Methods:** Duodenal biopsies were collected from 15 celiac disease patients on a strict long-term gluten-free diet (GFD) prior to and post a gluten challenge (PGC) and from 6 healthy control individuals (DC). Biopsy RNA was subjected to genome-wide 3’ RNA-Seq. Sequencing data was used to determine the differences between the three groups and was compared to sequencing data from the public repositories. The biopsies underwent morphometric analyses.

**Results:** In DC vs. GFD group comparisons, 167 differentially expressed genes were identified with 117 genes downregulated and 50 genes upregulated. In PGC vs. GFD group comparisons, 417 differentially expressed genes were identified with 195 genes downregulated and 222 genes upregulated. Celiac disease patients on a GFD were not “healthy”. In particular, genes encoding proteins for transporting small molecules were expressed less. In addition to the activation of immune response genes, a gluten challenge induced hyperactive intestinal wnt-signaling and consequent immature crypt gene expression resulting in less differentiated epithelium. Biopsy gene expression in response to a gluten challenge correlated with the extent of the histological damage. Regression models using only four gene transcripts described 97.2% of the mucosal morphology and 98.0% of the inflammatory changes observed.

**Conclusions:** Our gluten challenge trial design provided an opportunity to study the transition from health to disease. The results show that even on a strict GFD, despite being deemed healthy, patients reveal patterns of ongoing disease. Here, a transcriptomic regression model estimating the extent of gluten-induced duodenal mucosal injury is presented.

## Introduction

Celiac disease is an autoimmune disorder in which dietary cereal gluten is an essential inducer and driving force. The overall global prevalence of celiac disease is approximately 1%.^1^ There are differences between countries^1,2^, and at the population level the true prevalence has been shown to be increasing over time.^3^ During celiac disease pathogenesis, ingested gluten induces an autoimmune-like reaction in genetically susceptible subjects carrying the DQ2 or DQ8 human leukocyte antigen (HLA) haplotypes, which results in small intestinal villus atrophy and crypt hyperplasia and a wide array of gastrointestinal and non-gastrointestinal manifestations. In addition to dietary gluten and correct HLA genetics, the auto-antigen transglutaminase 2 (TG2), is needed.^4–6^ It was recently hypothesized that intestinal microbiota is a factor involved in the pathogenesis and clinical presentation of the disease.^7^ Currently, a lifelong gluten-free diet (GFD) is the only accepted treatment option for patients with celiac disease.

Ingestion of cereal gluten proteins is a prerequisite for small intestinal mucosal damage and inflammation in individuals carrying the HLA-DQ2 or HLA-DQ8 molecules. The HLA genes encoding the DQ2 and DQ8 molecules can be considered necessary but not sufficient factors, as only a small portion of the individuals with the correct HLA genetics will develop gluten-dependent celiac disease. Additionally, several non-HLA genes have been identified as contributing factors in celiac disease pathophysiology.^8^ Central to the disease pathogenesis are the abnormal intestinal T cell responses to gluten, and the ability of CD4^+^ T cells to recognize gluten epitopes is thought to be a driver of the disease.^9,10^ B cells, autoantibodies, and TG2, have also been shown to have an important function in celiac disease pathogenetic mechanisms.^9,11–14^ Despite the well-studied role of the immune system in celiac disease development, a complete picture of the pathophysiology, especially the early steps leading to the loss of gluten tolerance, is still missing.^13^

Intestinal epithelium, as one of the components in celiac disease development, has also been the focus of research. We believe that discovering the molecular mechanisms responsible for thwarting the differentiation program of the intestinal epithelial cells could lead to a better understanding of the mechanisms of celiac disease development and the consequent deterioration of the small intestinal mucosal morphology. Transcriptomic studies using microarray^15^ or RNA-Seq technologies^16–18^ and proteomic^19^ and epigenomic^20^ analyses were conducted on celiac disease patient biopsies. The majority of these studies used untreated patient samples, samples from patients on a GFD, and non-celiac disease samples as a control. Since mucosal damage is dose- and time-dependent,^21,22^ and since patients have different levels of gluten consumption at the time of their initial diagnoses, often large amount for decades, these confounding factors may lead to inconsistent results. To circumvent these shortcomings, we sought to exploit the gluten challenge study design^13,21–24^ and perform genome-wide transcriptomic analyses of celiac disease small intestinal biopsies from patients on a strict long-term GFD and compare the results to those of biopsies taken from the same celiac disease patients after a standard amount and time on a gluten challenge. We further compared these transcriptomic results with those obtained from non-celiac control patient biopsies. We also aimed to correlate the gene transcript levels with the degree of gluten-induced mucosal injury to create a molecular histomorphometry tool for assessing the extent of both duodenal mucosal architectural damage and inflammation.

## Materials and Methods

### Patients and biopsies

Fifteen adult HLA-DQ2 positive patients originally suspected for celiac disease due to symptoms, signs, and/or positive celiac serology who were also undergoing upper gastrointestinal endoscopies to receive a biopsy-proven diagnosis were enrolled for this study. All participants had followed a strict gluten-free diet for at least 1 year and for 16 years on average, and they were symptom free and negative for serum TG2 antibodies (GFD group). These patients were challenged, i.e. consumed 2—4 g of gluten daily for 10 weeks (post gluten challenge group, PGC). The control group included duodenal biopsies from six non-celiac individuals undergoing endoscopies due to dyspepsia or reflux symptoms. These patients had no detectable disease and were also negative for serum TG2 autoantibodies (non-celiac disease control group, DC). Before starting a gluten challenge, stool samples were collected to measure potential evidence of inadvertent gluten ingestion by using iVYLISA, a gluten-specific immunogenic peptide (GIP) assay, as described previously.^25–27^

Upper gastrointestinal endoscopies were performed, and duodenal forceps biopsies were immersed in PAXgene fixative and processed for paraffin block embedding (PaxFPE) using a standard formalin-free paraffin-infiltration protocol. For morphology, samples were stained with hematoxylin and eosin and measured using our validated morphometry rules for villus height (VH), crypt depth (CrD), and ratio (VH:CrD). To measure mucosal inflammation, CD3^+^ intraepithelial lymphocytes (IELs) were counted and reported as a density of positive T cells per 100 epithelial cells, as previously described.^24,28^

### RNA isolation and sequencing

Total RNA was extracted from the same duodenal PaxFPE biopsy specimens using further cuttings from where histomorphometry was to be measured and counted. For the extraction, an RNeasy Kit (Qiagen, Hilden, Germany) was used according to the manufacturer’s instructions. Library preparation and next-generation sequencing (NGS) was performed by Qiagen NGS Service. The library preparation was done using the QIAseq UPX 3’ Transcriptome Kit (Qiagen). A total of 10 ng of purified RNA was converted into cDNA NGS libraries, and a quality check was performed using TapeStation 4200 (Agilent, Santa Clara, CA) or Agilent Bioanalyzer (Agilent). Sequencing was performed on a NextSeq500 sequencing instrument according to the manufacturer’s instructions (Illumina, San Diego, CA). A secondary differential expression analysis involving normalization of unique molecular identifier (UMI) counts and a subsequent pairwise differential regulation analysis were done using DESeq2 package (Bioconductor).

### Alcian blue staining

Tissue sections were stained with 1% (w/v) Alcian blue 8GX (Sigma-Aldrich, Saint Louis, MO) in 3 % (v/v) acetic acid for 30 min at room temperature and washed in tap water. The sections were counterstained with Nuclear fast red (Sigma-Aldrich, Saint Louis, MO) for 3 min and dehydrated in an ascending series of ethanol, cleared in xylene, and mounted.

### Statistics and regression model

All statistical testing was performed using R: A Language and Environment for Statistical computing version 3.5.3 (Vienna, Austria) (Supplementary file with separately detailed regression models for mucosal morphology and inflammation). A Spearman’s rank correlation analysis was performed to examine the relationships between the differentially expressed genes and the VH:CrD or density of IELs. Regression models were created by the best subset selection approach. Among all possible combinations of genes from 1 to 6 fitted by a multiple linear regression, the best ones based on their cross validated prediction error, Bayesian information criterion (BIC), and adjusted R-square (Rsq) were selected. Models were evaluated by observed vs. predicted regression.^29^ As an additional validation step, principal component analysis (PCA) was performed on independent and publicly available dataset GSE134900.^30^ To quantify the separation between healthy (HC) and celiac disease (CD) groups in this independent dataset, the F-statistics metric was applied for four-dimensional data.^31^

### Ethics approval

The study was approved by the Ethics Committee of the Tampere University Hospital, Tampere, Finland (ETL-code R15185M). The patients gave their written consent to perform the study. The trial was registered at ClinicalTrial.gov, NCT02637141 and EudraCT 2015-003647-19.

All authors had access to the study data and reviewed and approved the final manuscript.

## Results

We analyzed the RNA from the same PaxFPE duodenal biopsy blocks where the VH, CrD, and VH:CrD were also measured. The morphology results (VH:CrD) of each study group are shown in Fig. 1A, and the grade of inflammation (IEL density) is shown in Fig. 1B. The morphometric results did not differ between the DC and GFD groups. Significant mucosal deterioration and increased inflammation was evident during a gluten challenge (GFD vs. PGC). Although the patients kept a strict GFD prior to the gluten challenge, the stool GIP results showed 4 out of the 15 had inadvertently ingested gluten just before stool sample collection (data not shown).

**Figure 1.**
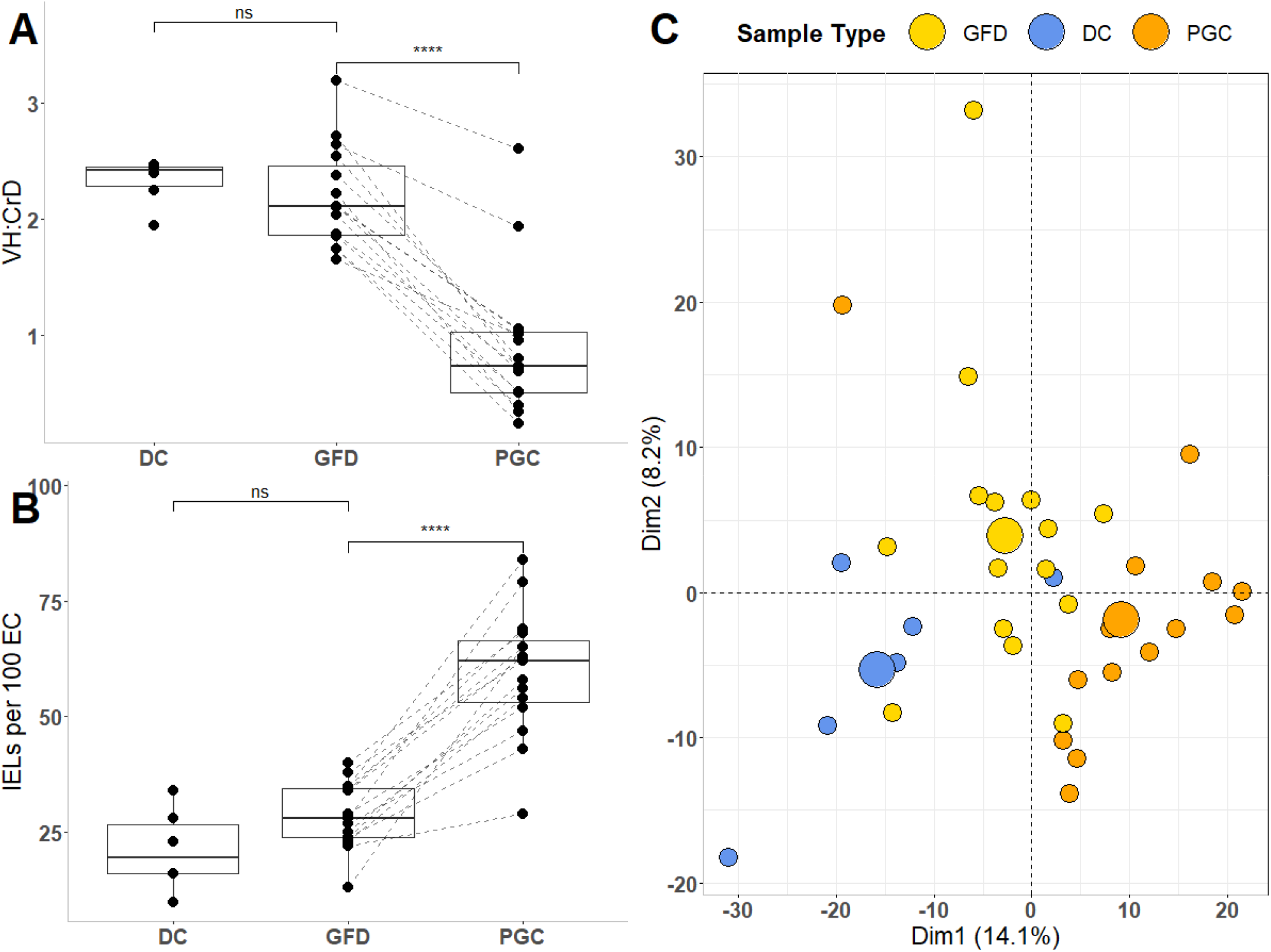
Histomorphometric data and principal component analysis plot based on transcriptome, from non-celiac disease control group, celiac disease patients on a gluten-free diet, and celiac disease patients post gluten challenge. A) Villus height and crypt depth ratio (VH:CrD) measurements. B) Counts of intraepithelial lymphocytes (IELs) per 100 enterocytes (EC). C) Principal component analysis (PCA) plot for all samples exploring biological differences between DC (blue), GFD (yellow), and PGC (orange) groups based on the gene expression profile with bigger spheres depicting the center of a distribution. Non-celiac disease control group (DC); gluten free-diet group (GFD); post gluten-challenge group (PGC).

### Genes affected by the gluten challenge

Biopsy-purified RNA was subjected to 3’ RNA-sequencing (RNA-seq). The unsupervised PCA plot for all samples indicated that the combination of dimensions (Dim1 and Dim2) explains 22.3% of the variance, showing a decent separation between the groups (Fig. 1C). When comparing RNA expression profiles from celiac disease patients in the GFD and PGC study groups, 417 differentially expressed genes were identified with 195 genes downregulated and 222 genes upregulated (FDR < 0.05 and |log2FC| ≥ 0.5). The top 100 differentially expressed genes, in a pairwise analysis of GFD vs. PGC, are shown in figures 2A and 2B (entire dataset in supplementary Fig. S1). Gene ontology analyses show that genes involved in cellular response to cytokines, including interferons, are over-represented (Fig. 2C). Similar results were obtained in previous RNA-seq studies where active celiac disease patients were compared to patients on a GFD or to non-celiac controls in a non-pairwise manner.^16,18^

**Figure 2.**
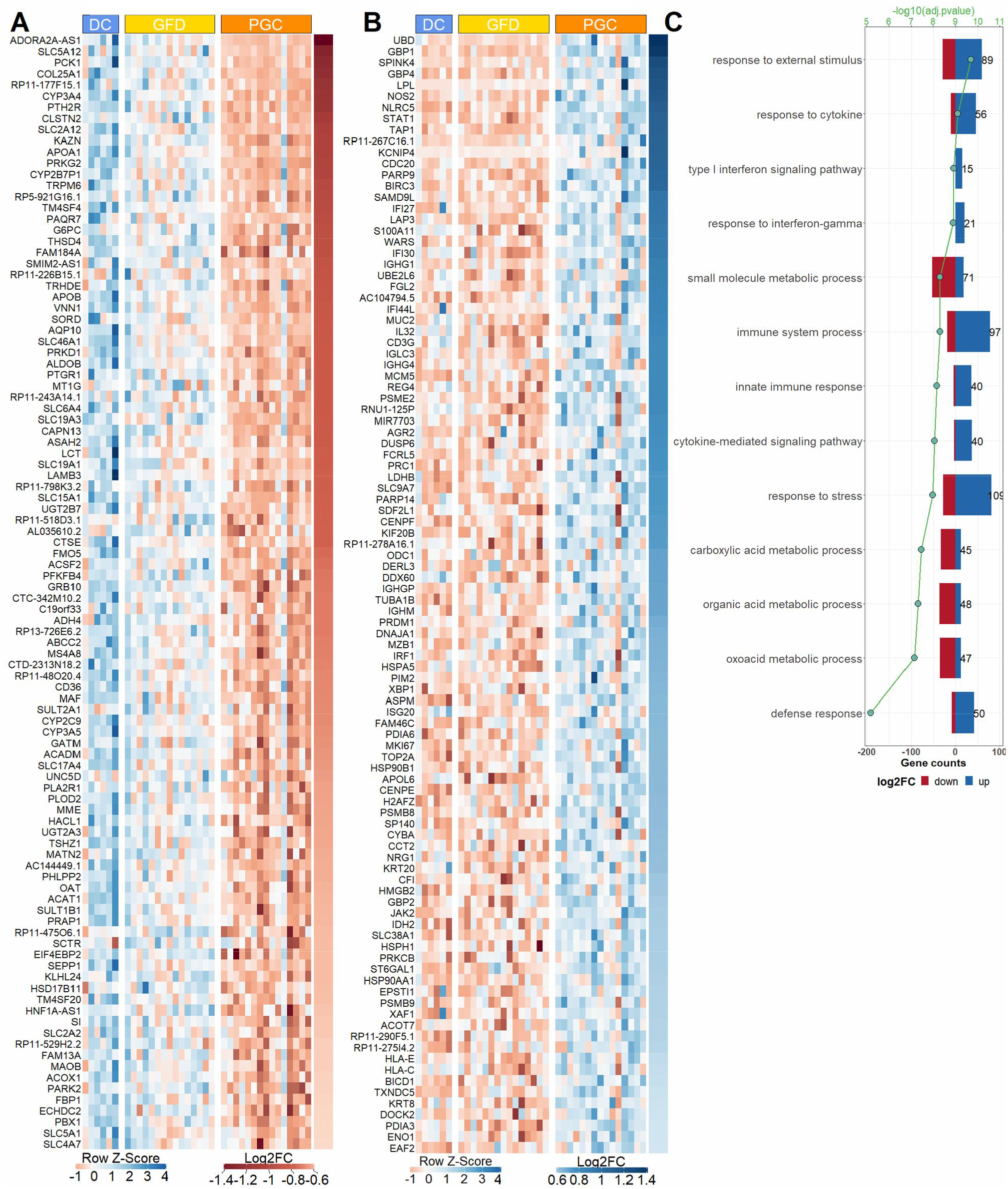
Differential intestinal mucosal gene transcriptome after 10 weeks of the gluten challenge in celiac disease patients. A) Heat map of the top 100 genes that are downregulated specifically after the gluten challenge. B) Heat map of the top 100 genes that are upregulated after the gluten challenge. C) Gene ontology (GO) terms enriched in identified set of genes (total 417 genes) specific for gluten-induced mucosal damage. Non-celiac disease control group (DC); gluten-free diet group (GFD); post gluten-challenge group (PGC).

### Genes affected by a gluten-free diet compared to non-celiac disease controls

As PCA, based on gene expression data (Fig. 1C), clearly discerned DC and GFD into separate groups, we analyzed the genes whose expression differ significantly already when celiac disease patients are on GFD when compared to the DC group. In total, 167 differentially expressed genes were identified (FDR < 0.05 and |log2FC| ≥ 0.5) with 117 genes downregulated and 50 genes upregulated in the GFD group when compared to the DC group (Fig. 3). Our results indicate that even though intestinal morphology in patients on a strict GFD is similar to that measured in DC patients (Fig. 1) based on gene transcription, the GFD group differs significantly from the DC group, showing a substantial number of differentially expressed genes. In general, the expression of these genes is not affected by the gluten challenge when comparing GFD and PGC groups. GO and Reactome pathway analyses reveal that patients on a GFD suffer from profoundly altered expression of genes with functions such as brush border assembly, developmental processes, transport of small molecules, and FOXO-mediated transcription of the cell cycle (Fig. 3C). For instance, within the transporter gene group, 24 genes are differentially expressed in the GFD group vs. the DC group. Of these, 22 are downregulated in the GFD group, including, for example, four transporter genes: FLVCR1 (heme transporter), SLC46A1 (folate and heme transporter), ATP2B1 (calcium transporter), and SLC39A4 (zinc transporter), as seen in Fig. 3D. This is in accordance with the clinical data, as it has been shown that patients on a GFD quite frequently suffer from micronutrient deficiencies.^32^ A list of the 24 transporter genes affected by a GFD is presented in Supplementary Table I.

**Figure 3.**
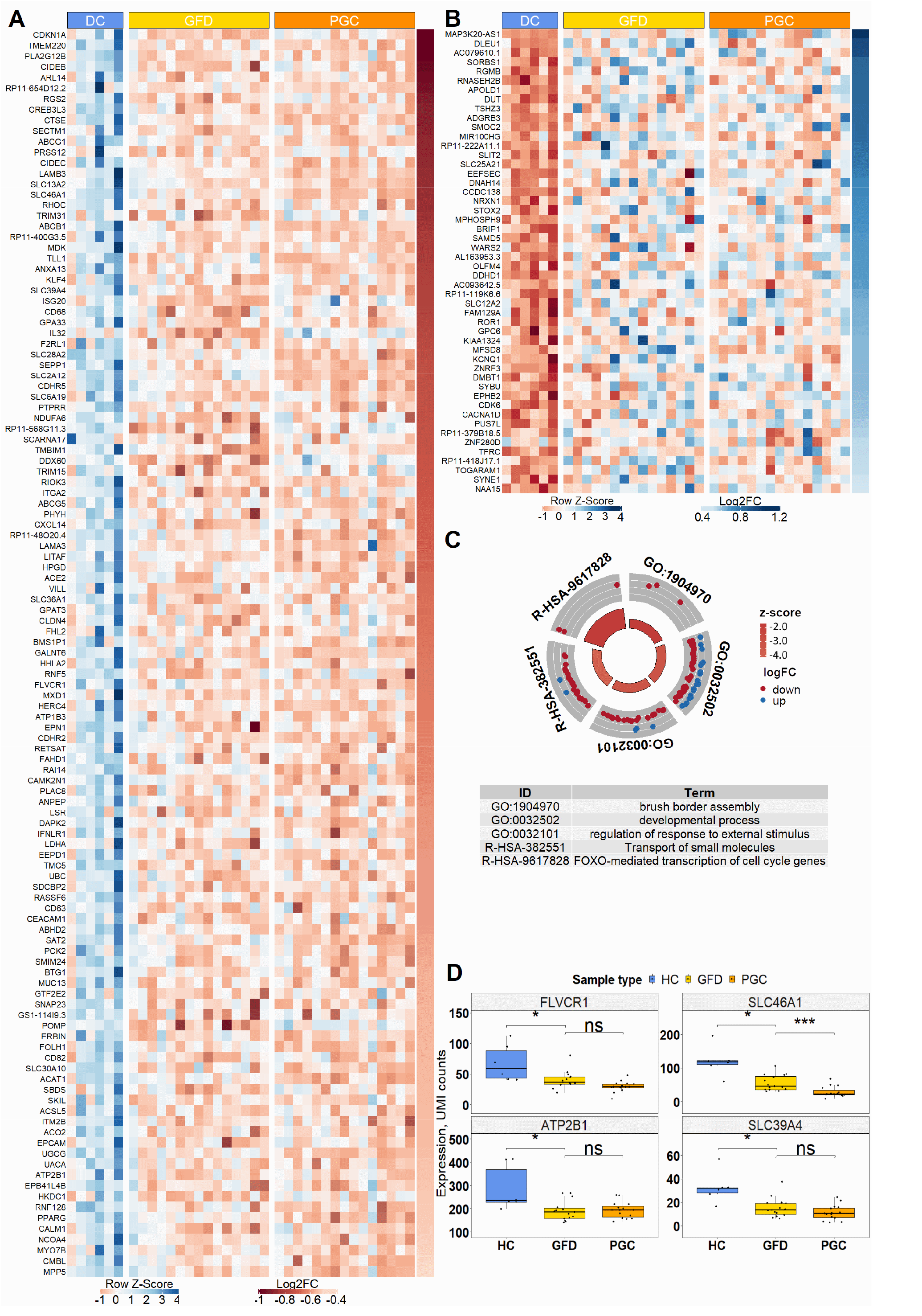
Differential intestinal mucosal gene transcriptome between healthy disease controls and celiac patients on a GFD. A) Heat map of 117 genes showing significantly lower expression in celiac disease patients in both GFD and PGC groups vs. the DC group. B) Heat map of 50 genes upregulated in both GFD and PGC groups vs. the DC group. C) Gene ontology terms and reactome pathways enriched in an identified set of genes (total 167) showing altered expression in the GFD group vs. the DC group. D) Changes of expression in some genes involved in the enriched “Transport of small molecules” Reactome pathway. Non-celiac disease control group (DC); gluten-free diet group (GFD); post gluten-challenge group (PGC).

### Hyperactive intestinal wnt-signaling is prominent in celiac disease patients after a gluten challenge but is also already enhanced during a gluten-free diet

It has been suggested that wnt-signaling is involved in celiac disease associated villous atrophy and crypt hyperplasia. More specifically, the occurrence of nuclear beta-catenin expressing cells is more prominent in active celiac disease patients compared to patients on a GFD and healthy controls.^33^ Moreover, it has been shown that active wnt-signaling compartments in the crypts of active celiac patients are expanded when wnt-target genes LGR5 and Sox9 are used as markers.^34^ We took advantage of our human RNA-Seq data by comparing it with our previous data in which intestinal differentiation was induced in vitro in mouse intestinal organoids with wnt-inhibitor IWP2 and gene expression was analyzed with Gro-Seq before and after differentiation.^35^ These data comparisons enable us to characterize the wnt-mediated component of mucosal damage in celiac disease. Out of all 417 differentially expressed genes after the gluten challenge, 90 (21.6%) belong to the wnt-component (Fig. 4A). An approximately similar size of wnt-component is also affected when celiac patients on a GFD are compared to non-celiac disease control patients, as 42 genes out of all 167 differentially expressed genes (25.1%) belong to the functional wnt-component in intestinal organoids (Fig. 4A). To address the hypothesis that wnt-signaling is pathogenically hyperactive in celiac disease, we analyzed the directionality of the gene expression, and indeed found this to be the case. Of the 195 genes repressed by the gluten challenge, 48 (24.6%) belong to the group of genes induced by wnt-inhibition (Fig. 4B). Reciprocally, only 5.9% (13 out of 222) of the genes upregulated by the gluten challenge are also induced in wnt-inhibition (Fig. 4C). Reactome pathway analysis reveals that these 13 oppositely responding genes belong in the immune response category (Fig. S3), which was specifically induced in the PGC group (Fig. 4F). Interestingly, when comparing celiac disease patients on a GFD to DC patients, a similar kind of over-active wnt-component is also evident, as 28.2% (33 out of 117) of genes downregulated in the GFD group compared to the DC group belong to the wnt-inhibition induced wnt-component (Fig. 4B). On the other hand, only 2% (1 out of 50) of genes upregulated by the gluten challenge belong to the wnt-inhibition induced wnt-component (Fig. 4C). Moreover, a similar trend can be seen in genes downregulated by wnt-inhibition, as 13.1% (29 out of 222) of genes upregulated by a gluten challenge belong to the wnt-component repressed by wnt-inhibition (Fig. 4D). Correspondingly, not a single gene of the downregulated gene group affected by the gluten challenge falls into the category of wnt-component repressed by wnt-inhibition (Fig. 4E). To conclude, our data suggest that in active celiac disease, in addition to activation of immune response genes, wnt-signaling is conspicuously overactive in the small intestine, favoring immature crypt gene expression at the expense of less differentiated epithelium. Interestingly, this pathogenic “wnt-on” state, excluding the inflammatory component, seems to already be active in celiac patients on a GFD.

**Figure 4.**
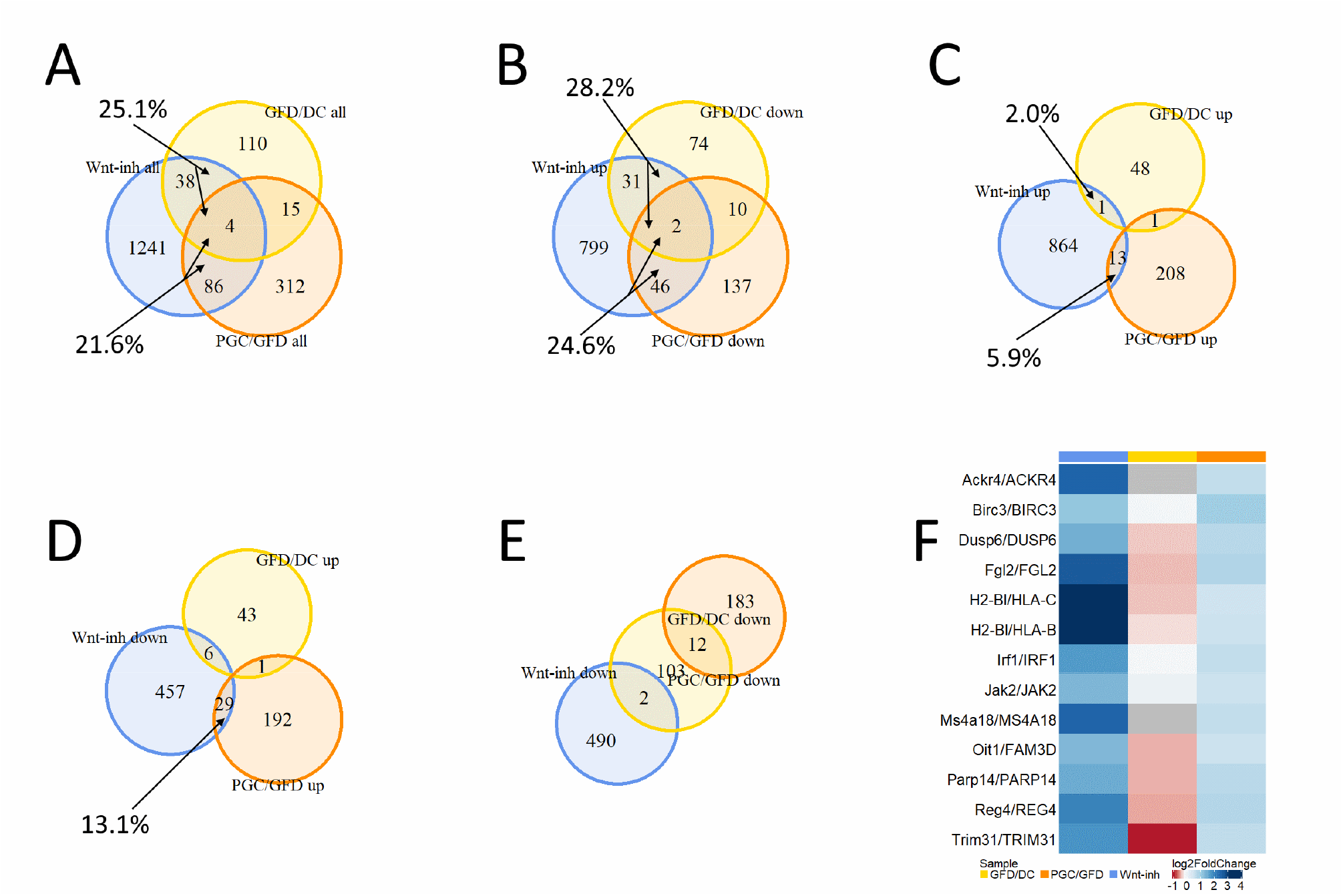
Hyperactive wnt-component in celiac disease. Venn diagrams of A) all differentially expressed genes in intestinal organoids after wnt-inhibition (blue spheres) and GFD/DC (yellow spheres) and PGC/GFD (orange spheres) comparisons. B) Upregulated genes after wnt-inhibition and downregulated genes indicated by comparisons. C) Upregulated genes after wnt-inhibition and upregulated genes indicated by comparisons. D) Downregulated genes after wnt-inhibition and upregulated genes indicated by comparisons. E) Downregulated genes after wnt-inhibition and downregulated genes indicated by comparisons. F) The immune response category genes that are specifically induced only in the PGC. Non-celiac disease control group (DC); gluten-free diet group (GFD); post gluten-challenge group (PGC).

### A gluten challenge induces a secretory cell type signature and inhibits absorptive enterocyte maturation

Increased epithelial proliferation and reduction of the differentiated cells and consequent crypt hyperplasia and villous atrophy are the hallmarks in the gluten-dependent manifestation of the small intestinal mucosal lesion.^4^ Differentiation of both absorptive and secretory epithelial cells has been shown to be affected in celiac disease.^36,37^ We compared our biopsy transcriptomic data to published data on single cell RNA-Seq (scRNA-seq) studies on the small intestinal epithelium.^38^ We found that, congruent with the previous studies,^39^ mature absorptive enterocyte lineage genes were almost unanimously downregulated in PGC patients. Interestingly, however, we found that genes for secretory cell signatures were upregulated during the gluten challenge (Fig. 5A), and most of these genes belong to the goblet cell signature. However, the number of goblet cells per mm of epithelium was not significantly different between the groups (Figs. 5B and 5C).

**Figure 5.**
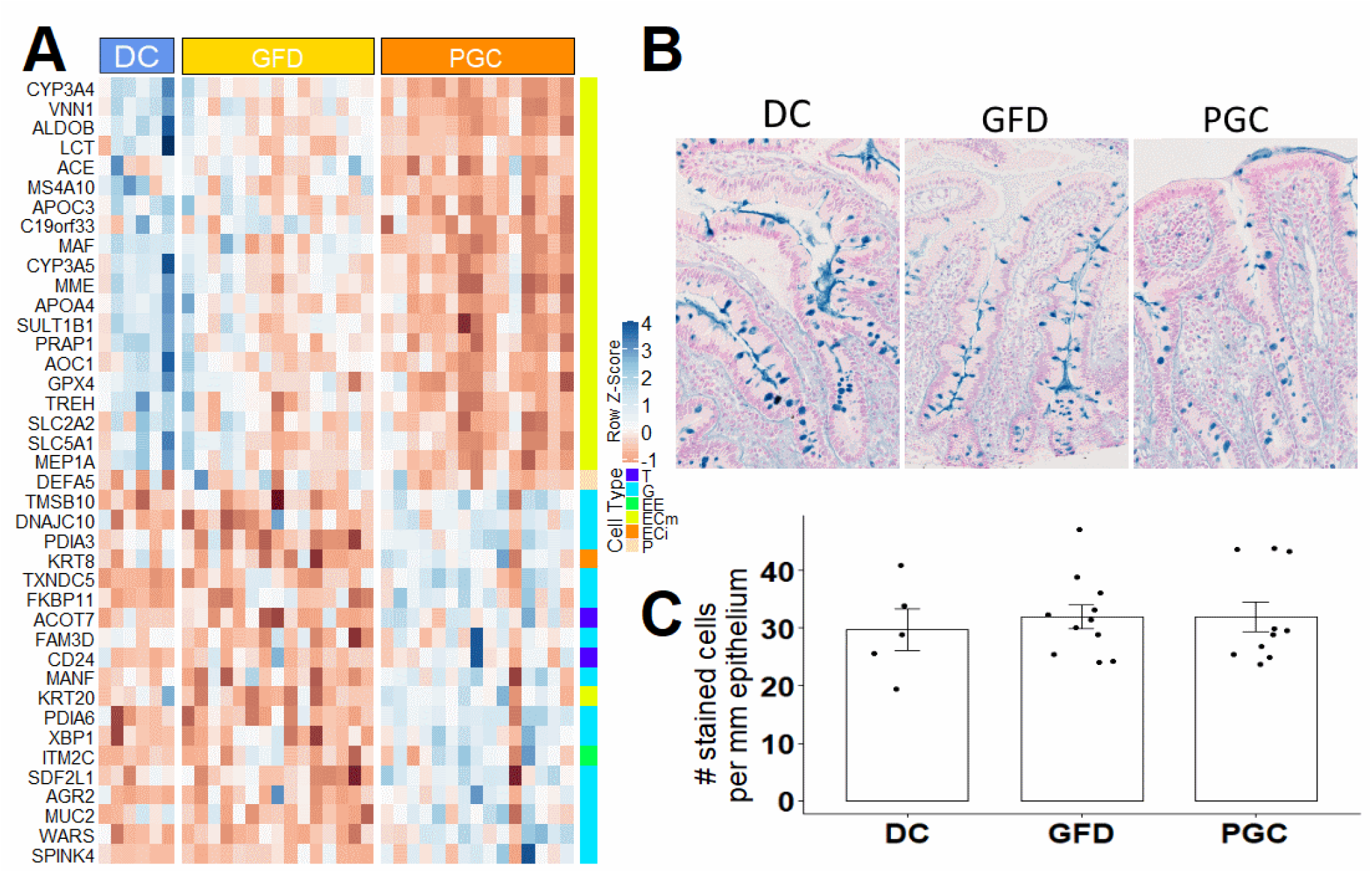
Dietary gluten inhibits absorptive cell specific gene expression and evokes secretory cell type gene expression. A) Gene expression heat map of the intestinal epithelial cell type specific genes. Tuft cells (T); goblet cells (G); enteroendocrine cells (EE); enterocyte mature (ECm); enterocyte immature (ECi); paneth cells (P). B) Alcian Blue goblet cell staining in DC, GFD, and PGC patients at 40x magnification. C) Number of goblet cells per mm of epithelium. Non-celiac disease control group (DC); gluten-free diet group (GFD); post gluten-challenge group (PGC).

### Morphometric regression models and validation in gluten-induced duodenal mucosal injury

All identified 112 differentially expressed protein-coding genes that significantly correlate with VH:CrD (Spearman’s rank, |rho|> 0.65), and all 98 differentially expressed protein-coding genes that significantly correlate with IEL densities (Spearman’s rank, |rho|> 0.6) were selected for molecular morphometry model creation. They were then separately analyzed for duodenal mucosal morphology (VH:CrD) and inflammation (IEL density). The gene expression correlations with the extent of gluten-induced histological damage and inflammation are presented as heat maps in supplementary Fig. S2. The subsequent best subset selection approach (Supplementary materials and methods) computationally reduced the number of genes, which describes VH:CrD (Model 1) and IEL density (Model 2) changes to four genes for each model (Table I). Created models describe 97.2% of observed VH:CrD and 98.0% of IEL number variabilities, and there is a strong correlation between the predicted and observed ratios (Figs. 6A and 6B). For the validation of our models’ applicability to independent data, the PCA was performed on dataset GSE134900^30^ (Figs. 6C-E). F-statistics suggest that by using the expression of four genes predicted by our models to describe the variance in histomorphology parameters we are able to discriminate between healthy and celiac disease patients better than by using the expression of four randomly selected genes (F = 78 and 36 for VH:CrD and IEL density, respectively, compared to F = 10 for randomly selected genes).

**Table I.**
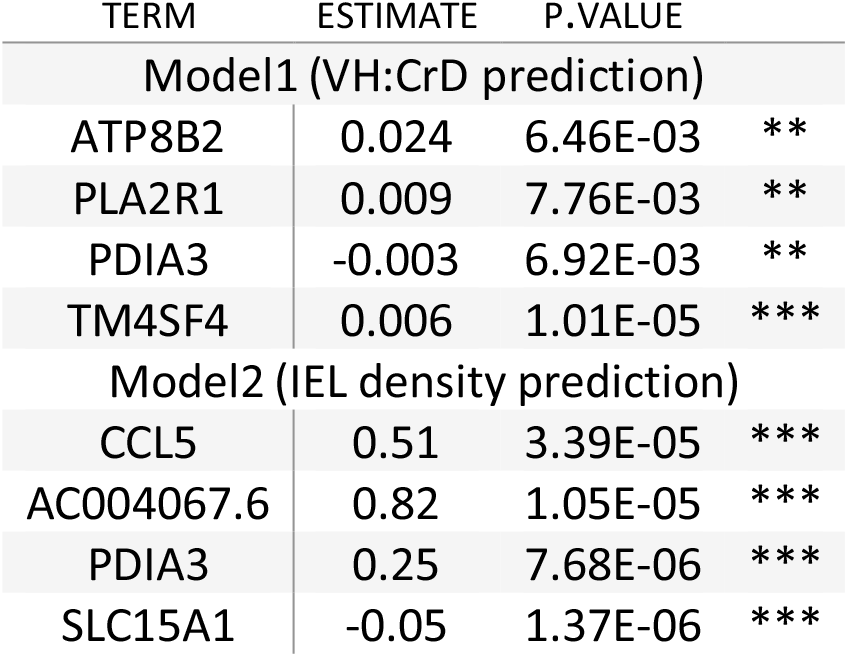
Model 1, villus height crypt depth ratio (VH:CrD) and Model 2 CD3^+^ intraepithelial (IEL) density coefficients

| TERM | ESTIMATE | P.VALUE |  |
| --- | --- | --- | --- |
| Model1 (VH:CrD prediction) |  |  |  |
| ATP8B2 | 0.024 | 6.46E-03 | ** |
| PLA2R1 | 0.009 | 7.76E-03 | ** |
| PDIA3 | -0.003 | 6.92E-03 | ** |
| TM4SF4 | 0.006 | 1.01E-05 | *** |
| Model2 (IEL density prediction) |  |  |  |
| CCL5 | 0.51 | 3.39E-05 | *** |
| AC004067.6 | 0.82 | 1.05E-05 | *** |
| PDIA3 | 0.25 | 7.68E-06 | *** |
| SLC15A1 | -0.05 | 1.37E-06 | *** |

**Figure 6.**
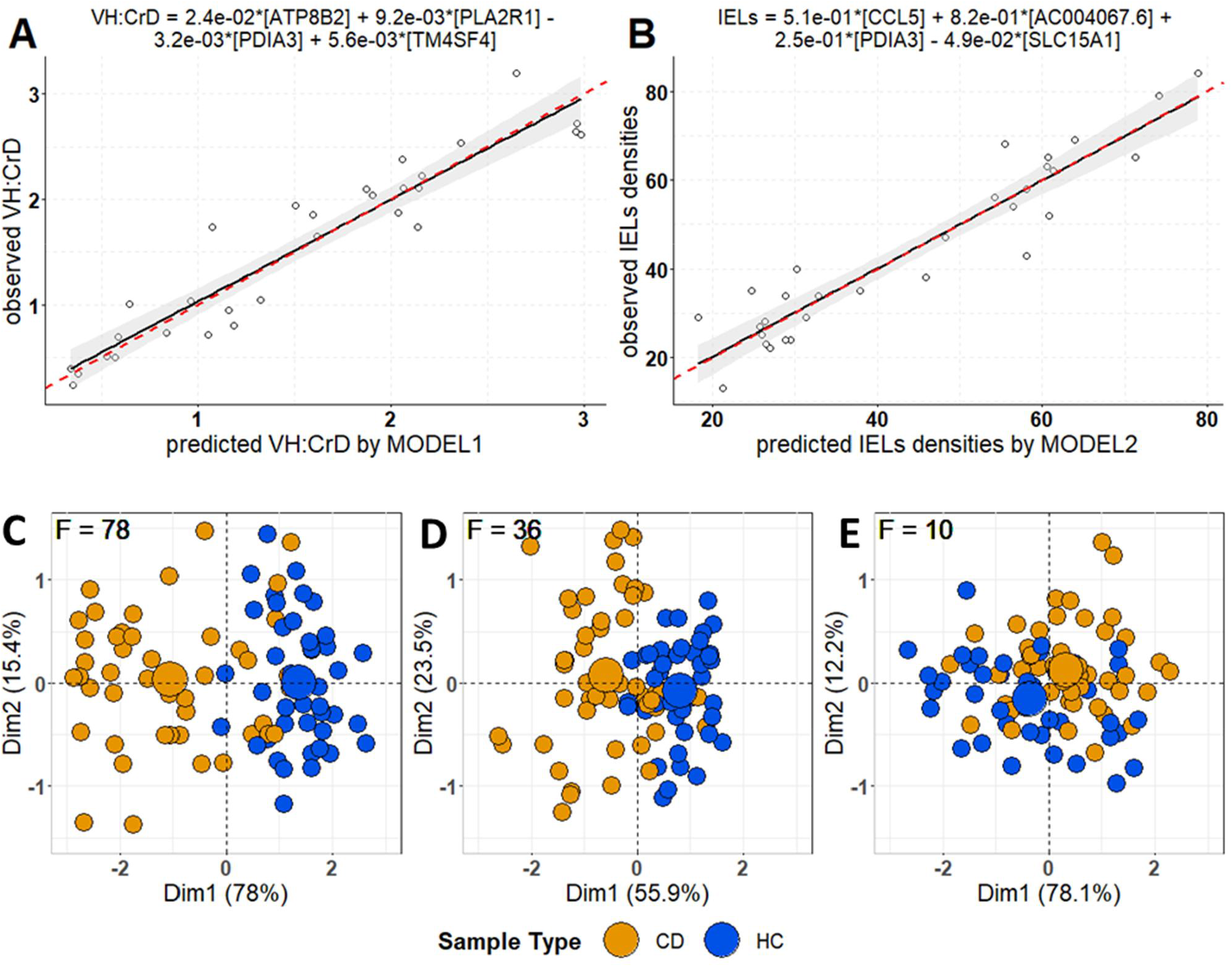
Observed vs. predicted regression scatter plots for Model 1 predicting villus height and crypt depth ratio (VH:CrD) (A) and for Model 2 predicting density of intraepithelial lymphocytes (IELs) (B). Epithelial cells (ECs). Red dashed line represents ideal regression case, where x = y. C-E) For validation, a graphical representation of principal component analyses (PCA, Dim1, and Dim2) applied to expression of four genes involved in Model 1 (C), Model 2 (D), and four other random genes (E) selected out of the pool of the all detected genes extracted from publicly available dataset GSE134900 is shown. Celiac disease (CD); healthy control (HC); F-statistics (F).

## Discussion

To our understanding, this is the first genome-wide RNA-Seq study with paired duodenal biopsies from celiac disease patients on strict long-term gluten-free diets before and after 10 weeks of a gluten challenge. We strived to pinpoint not only the early gluten-dependent gene transcripts but also genes that are constantly differentially expressed in celiac disease patients vs. control patients. The present results show that at the molecular level, healthy intestinal functions were not reinstated upon long-term GFD restrictions. Our novel finding is that genes, particularly those encoding proteins for transporting small molecules, were significantly less expressed in patients on a GFD compared to healthy individuals, which can contribute to micronutrient deficiency. For example, we show downregulation of two heme transporter expressions (FLVCR1 and SLC46A1) and upregulation of Transferrin receptor (TFRC), the gene responsible for iron delivery from transferrin to the cell. The TFRC gene is also known to mediate the IgA-mediated gliadin transport during celiac disease pathogenesis.^40,41^ Related to folate deficiency, we found significant downregulation of the gene SLC46A1, which encodes a folate transporter. Linked to calcium and zinc deficiency, genes involved in Ca^2+^ homeostasis (ATP2B1 and CALM1) and the zinc transporter SLC39A4 showed decreased levels of expression in patients on a GFD compared to healthy controls. In fact, micronutrient deficiencies are relatively common in celiac disease patients, even those who are on a strict GFD for long periods. For example, iron, folate, and B12 along with mineral and vitamin D deficiencies are frequent.^27,42^ Gluten-free products may not be of optimal nutritional quality^43^, and lack of fortification may increase the risk of the deficiencies in celiac sufferers.^44^ Kreutz et al. speculated that, in addition to inadequate dietary intake, diminished uptake due to intestinal dysfunction would contribute to these nutrient deficiencies in celiac disease patients on a GFD.^42^ Our data clearly indicate that the low expression of micronutrient transporters in the intestine may be the cause for the deficiencies observed in clinics. We further believe our transcriptomic findings are due to inadvertent gluten ingestion and indication of disease due to early mucosal injury. In fact, in the inadvertent gluten ingestion, while still following a strict GFD, was evidenced to occur in 27% of the present patient population. Complete elimination of dietary gluten seems impossible to maintain due to issues such as contamination of gluten-free foods, resulting in the ingestion of hundreds of milligrams of gluten per day in an attempted gluten-free diet.^27,45,46^ Inadvertent gluten ingestion is clinically difficult to detect, as serum TG2 or endomysial antibodies do not pick up most of the patients with persistent villous atrophy on gluten-free diets.^47^ When the stool GIPs measure gluten ingestion the day or so before testing, the duodenal mucosa is reacting upon long-term and significant inadvertent gluten ingestion. Recently, quantitative histology with well-oriented biopsy sections was shown to reveal villus atrophy in the majority of patients with celiac disease compliant with well-controlled gluten-free diets.^48^ The celiac disease patients in the present study did not reach a duodenal mucosal healing described as fully normal.^49^ Even if our six control patients without evidence of celiac disease had morphologically similar small intestinal mucosal findings, they clearly differed from celiac disease patients on a GFD as related to the transcriptomic findings. Furthermore, our rather small control patient group had a reason for endoscopy and was, in fact, comprised of disease control patients, which may mean they were not fully “normal.” When the duodenal mucosa in celiac disease evidence “normalization” upon a GFD, there is still disease at a molecular and even clinical level. It is intriguing to speculate that transcriptomic changes such as those seen in the present study in celiac disease patients on a strict GFD could also occur in latent celiac disease i.e., patients with existing gluten-dependent disease where the disease is not yet manifested at the mucosal level. Such patients “excluded on biopsy” for celiac disease have been shown to have micronutrient deficiencies and even bone disease with fractures, and their duodenal mucosa may deteriorate to a state typical for celiac disease only later, which includes crypt hyperplastic lesion with villous atrophy.^50–53^

Our pairwise analysis indicates that genes specific for absorptive enterocytes were almost unanimously downregulated in celiac disease patients after a gluten challenge. This has also been shown in other studies where active celiac disease patients were compared to healthy controls or celiac patients on a GFD in a non-pairwise manner.^36,37,39^ Deficient enterocyte differentiation has been suggested to be ascribed to hyperactive wnt-signaling in the disease, as wnt-signaling needs to be inhibited for enterocyte differentiation to occur.^49^ In fact, the number of cells expressing nuclear beta-catenin in the crypt, as a proxy for active wnt-signaling, has been shown to be increased along with expression of a few studied wnt-target genes in active celiac disease^33,34^. We addressed this by comparing the RNA-seq gene expression data from patients to our wnt-component gene expression data from intestinal organoids.^35^ Our data shows that in active celiac disease wnt-signaling is clearly hyperactive, which is most probably causing the compromised absorptive cell differentiation. In addition, this pathogenic hyperactive wnt-signaling state (excluding the inflammatory component) seems to already be active in celiac disease patients on a GFD. In fact, contrary to compromised absorptive cell differentiation, we saw that secretory cell type gene expression is induced after a gluten challenge. The majority of upregulated secretory cell line markers in our datasets belong to goblet cells. A reduction in the number of goblet cells has been reported in active celiac disease^52,53^, but counterintuitively some of the goblet cell markers, including SPINK4^54^ and MUC2^55^, are upregulated. Moreover, in vitro studies with organoids have shown that intestinal organoids derived from celiac disease patients are more poised for secretory cell differentiation. For example, MUC2 was expressed more in organoids derived from active celiac disease patients compared to organoids derived from non-celiac individuals.^37^ Our genome-wide in vivo data shows that secretory cell markers, mainly those of the goblet cells, are consistently upregulated in celiac disease after a gluten challenge. Contrary to absorptive cell differentiation, Notch-signaling needs to be off and wnt-signaling needs to be on for secretory cell differentiation to take place.^49^ Since it is known that proinflammatory cytokines can induce genes involved in the immune response in intestinal enterocytes,^56^ one may argue that these cytokines are the primary cause for the upregulation of secretory cell markers in active celiac disease. Th1 and Th2 inflammatory cytokines are known to induce goblet cell differentiation through activation of the PI3K/Akt pathways.^57,58^ Celiac disease in its active phase is characterized by activation of the Th1 gene expression profile.^59^ TNF-α, IL-4, and IL-13 have been shown to induce the expression of MUC2 via NF-kB mediated by mitogen-activated protein kinase pathways in vitro.^60,61^ There is evidence that over-production of MUC2 in inflammatory bowel disease depletes goblet cells. In vitro studies showed that mucus hypersecretion caused severe endoplasmic reticulum stress and apoptosis of goblet cells.^62^ This could possibly explain our finding that in celiac disease after a gluten challenge the expression of MUC2 and other goblet cell markers are increased without any changes in goblet cell numbers. To conclude, in active celiac disease, hyperactive wnt-signaling seems to impede absorptive cell differentiation, but at the same time, together with inflammatory cytokines, it might poise secretory cell differentiation.

The basis for a celiac disease diagnosis is the characteristic gluten-triggered and dependent small intestinal mucosal lesion (villous atrophy with crypt hyperplasia), which is most often associated with inflammation seen as an increased intraepithelial T cell density. Traditional celiac disease diagnostics based on grouped classifications (so called Marsh-Oberhuber classes) are highly subjective, and interobserver reproducibility between pathology readers is not optimal.^63–65^ The statement by Picarelli et al., where they stressed the limits of histological interpretation due to a lack of uniformity in the use of Marsh-Oberhuber classification,^66^ is still relevant as evidenced by a recent 33-celiac center study where one reader evaluated the mucosa to be Marsh 0 and the other graded it from Marsh 1 to 3c and vice versa (supplementary Table S20 Werkstetter et al.).^67^ In the present study, we adopted our standard operating procedures and validated quantitative morphometry separately for biopsy morphology and inflammation to measure the continuum of gluten-induced mucosal injury in celiac disease.^13,21,24,28^ These methods were also adopted in recent clinical drug/vaccine trials.^22,23,48^ In the present study, we show that the expression of a selected subset of protein-coding genes correlates extremely well with the extent of morphological damage and inflammation in control patients and celiac disease patients on a GFD before and after the gluten challenge (presented as heat maps in the order of VH:CrD and IEL density, supplementary figure S2). It should be noted that the RNA was extracted from separate cuttings but from the same paraffin-embedded biopsy blocks that had been used for morphometry measurements, thus avoiding the patchy lesion pitfalls observed between biopsies in short gluten challenges using low and moderate amounts of gluten. Gene expression seems to determine the state of a tissue, and the changes in expression are associated with the severity of the injury. We have already, as a proof of concept, moved toward “molecular histomorphometry” using a candidate gene approach.^24^ Also, other attempts to build a molecular model for grouped mucosal injury classification have been presented.^18,71^ In our study, we modeled the behavior of continuous metric parameters (VH:CrD and the density of CD3^+^ IELs), which were assigned to each endoscopic biopsy, and a multiple regression analysis was applied. Thus, we first predicted gluten-induced mucosal damage expressed as VH:CrD and IEL parameter changes by studying the changes in gene expression. Additionally, the whole mass of transcriptomic data obtained after RNA-sequencing (approximately 19,000 gene transcripts) was used for predictor selections. In conclusion, we show that by using transcriptomic data one may predict histomorphological parameters with a high accuracy. In fact, the subset selection approach computationally reduced the number of genes to four for both the morphology and inflammation predictions. The created models described 97.2% of observed VH:CrD and 98.0% of the IEL densities; we show a strong correlation between the predicted and observed ratios. We foresee this approach to be exploited in celiac disease diagnostics and in drug trials monitoring the efficacy of novel treatments focused on circumventing subjective biopsy readings.

We acknowledge the present study limitations. The present results are drawn from a rather small patient group. Also, our designed molecular morphometry models are based on RNA-sequencing data, and additional validation with RT-qPCR is warranted in a separate clinical gluten challenge study. On the other hand, we provide genome-wide duodenal biopsy transcriptomic data from a human source where gene expression correlated with the extent of histological injury continuum studied in control patients and celiac disease patients on a strict GFD and after a gluten challenge. It should be noted that we addressed only early gluten-induced transcriptomic changes in respect to morphology and inflammation and did not study, for example, newly diagnosed celiac disease patients who had ingested 10—20 g of gluten for decades. However, we were able to validate our data with the published independent data (dataset GSE134900)^30^, suggesting that our regression model could be applicable for newly diagnosed celiac disease patients.

To conclude, we provide transcriptomic resource data not only to determine gluten-dependent morphology and inflammation in celiac disease but also to pinpoint genes potentially important in pathogenesis, in finding genes for novel drug targets, and for biomarker/surrogate marker research. We further show that even a long and strict gluten-free diet is not sufficient to abate relatively common micronutrient deficiencies among celiac disease patients. Adherence to a strict gluten-free diet can be onerous to maintain, and since contamination of gluten-free foods is common, complete elimination of dietary gluten is difficult, if not impossible, to attain. Our results describe aforementioned adverse conditions at the molecular level and warrant the pursuit of novel adjunct treatments for celiac disease.

## Data Availability

Transcript Profiling
Data series entry GSE145358 for the RNA-Seq data is accepted in GEO.

## Acknowledgments

We thank patients participating for making this study possible. We also thank the expert staff for their participation in the sample collection.

---

**Supplementary figure 1.**
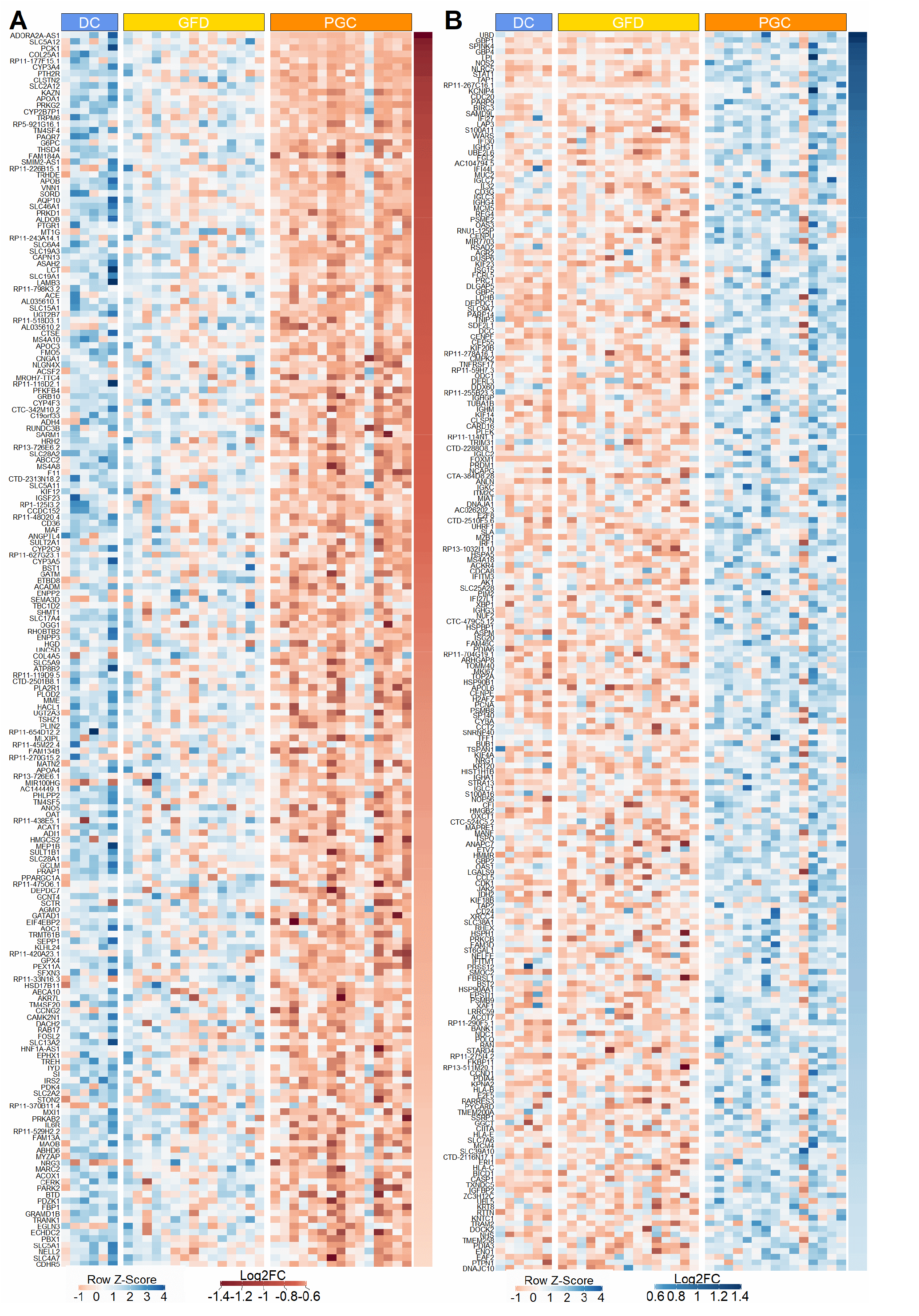
Differential intestinal mucosal gene transcriptome after 10 weeks gluten-challenge. A) Heat map of 195 genes that are downregulated specifically after gluten-challenge. B) Heat map of 222 genes that are upregulated after gluten-challenge. (FDR < 0.05 and |log2FC| ≥ 0.5).

**Supplementary Table I.**
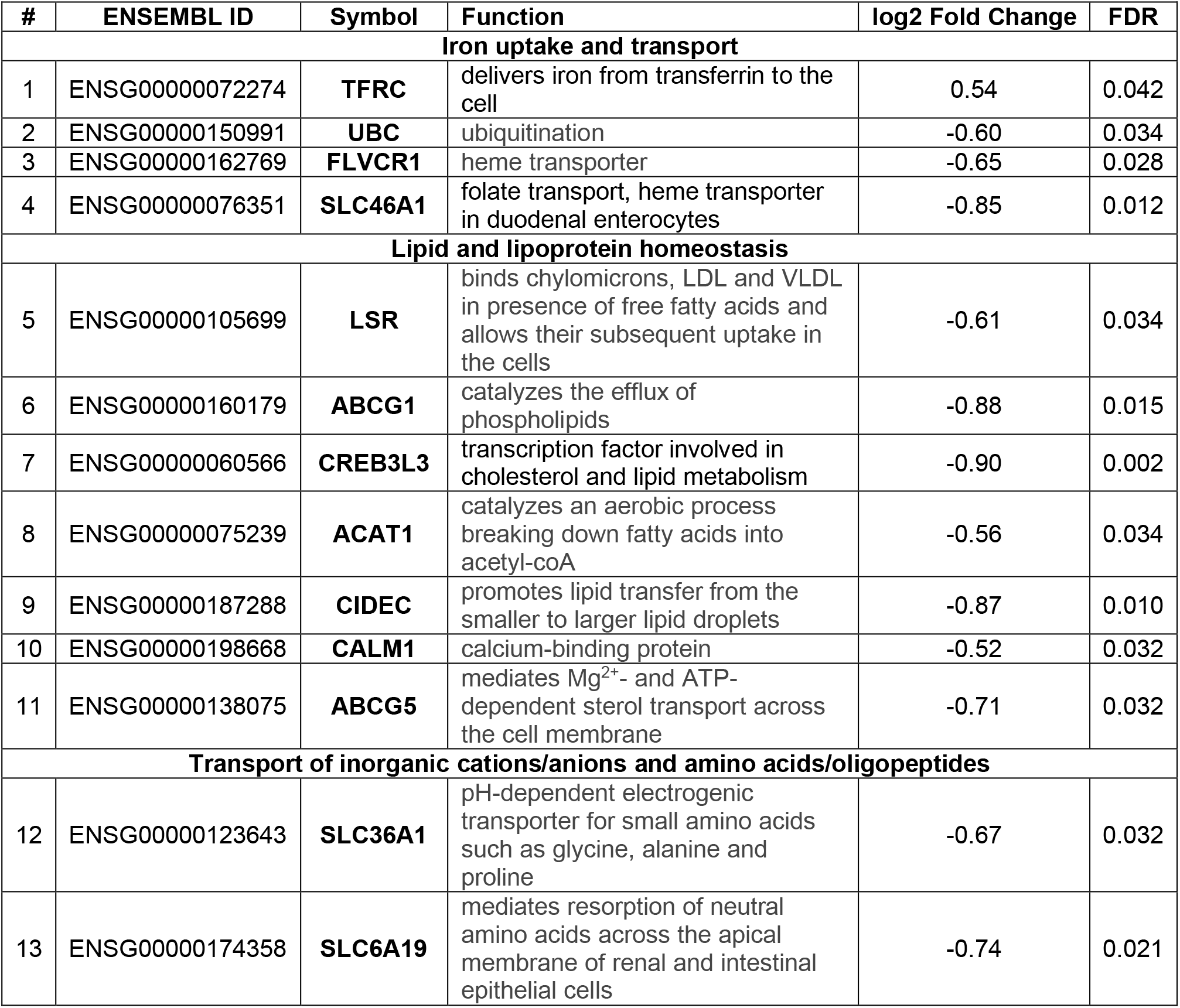
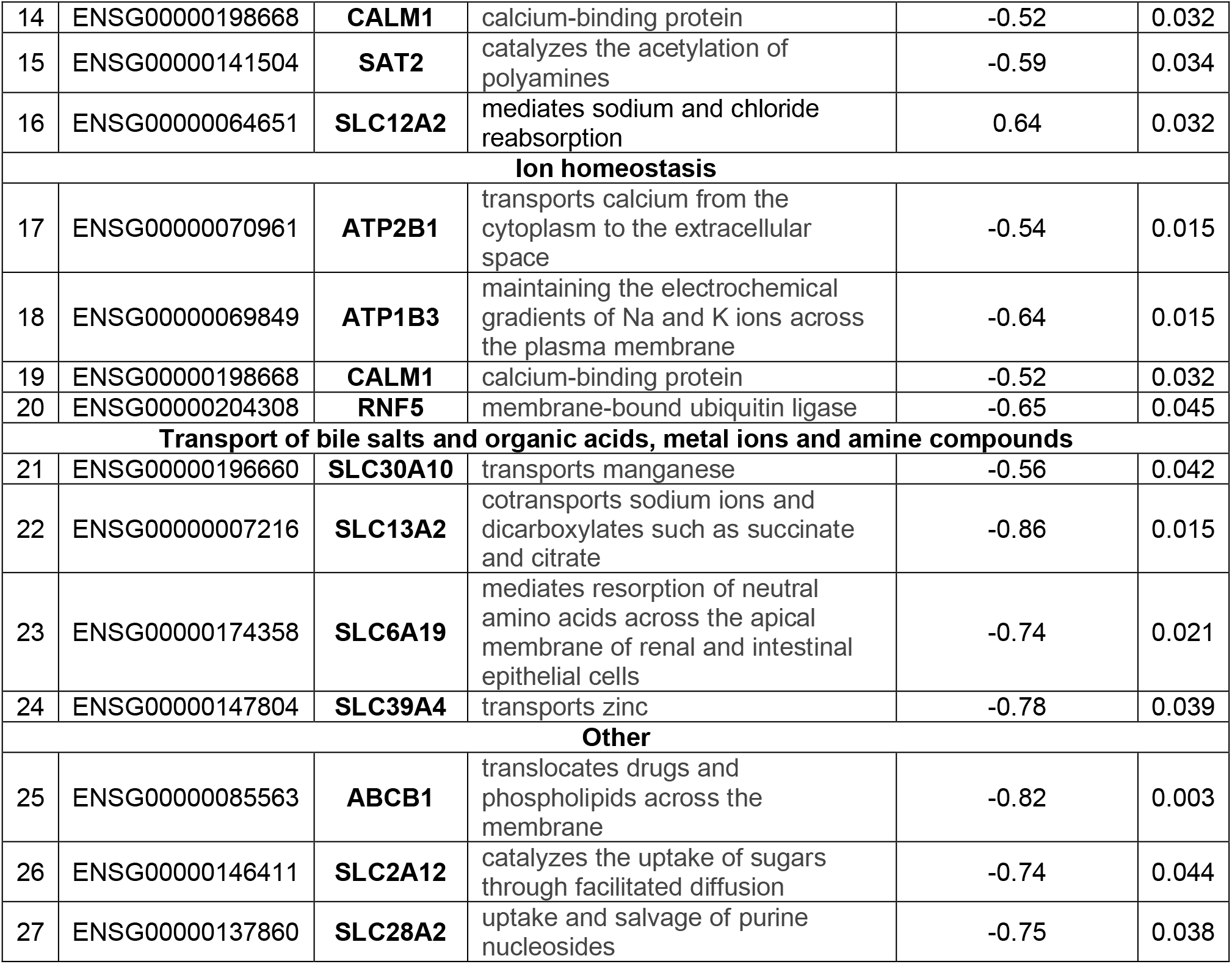
List of the 24 genes affected already in GFD within the group of transport of small molecules are grouped according to their specific function. Gene ID, gene symbol, function and numerical values for DC vs. GFD as log2 fold change and adjusted p-values are shown.

**Supplementary figure 2.**
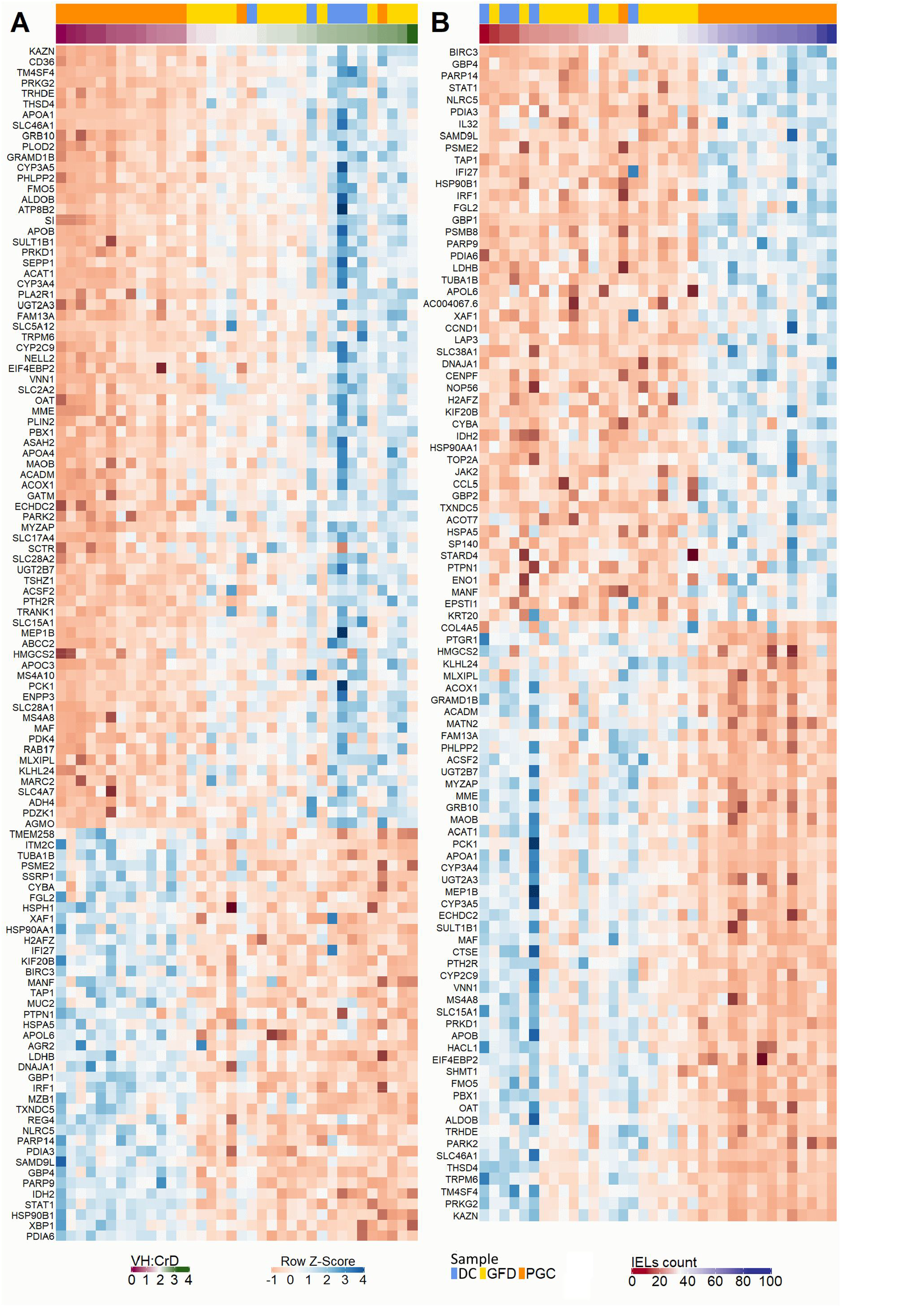
The gene expression correlations with the extent of gluten-induced histological damage, (Spearman’s rank, |rho|> 0.65) (A) and inflammation, (Spearman’s rank, |rho|> 0.6) (B) are presented as heat maps.

**Supplementary figure 3.**
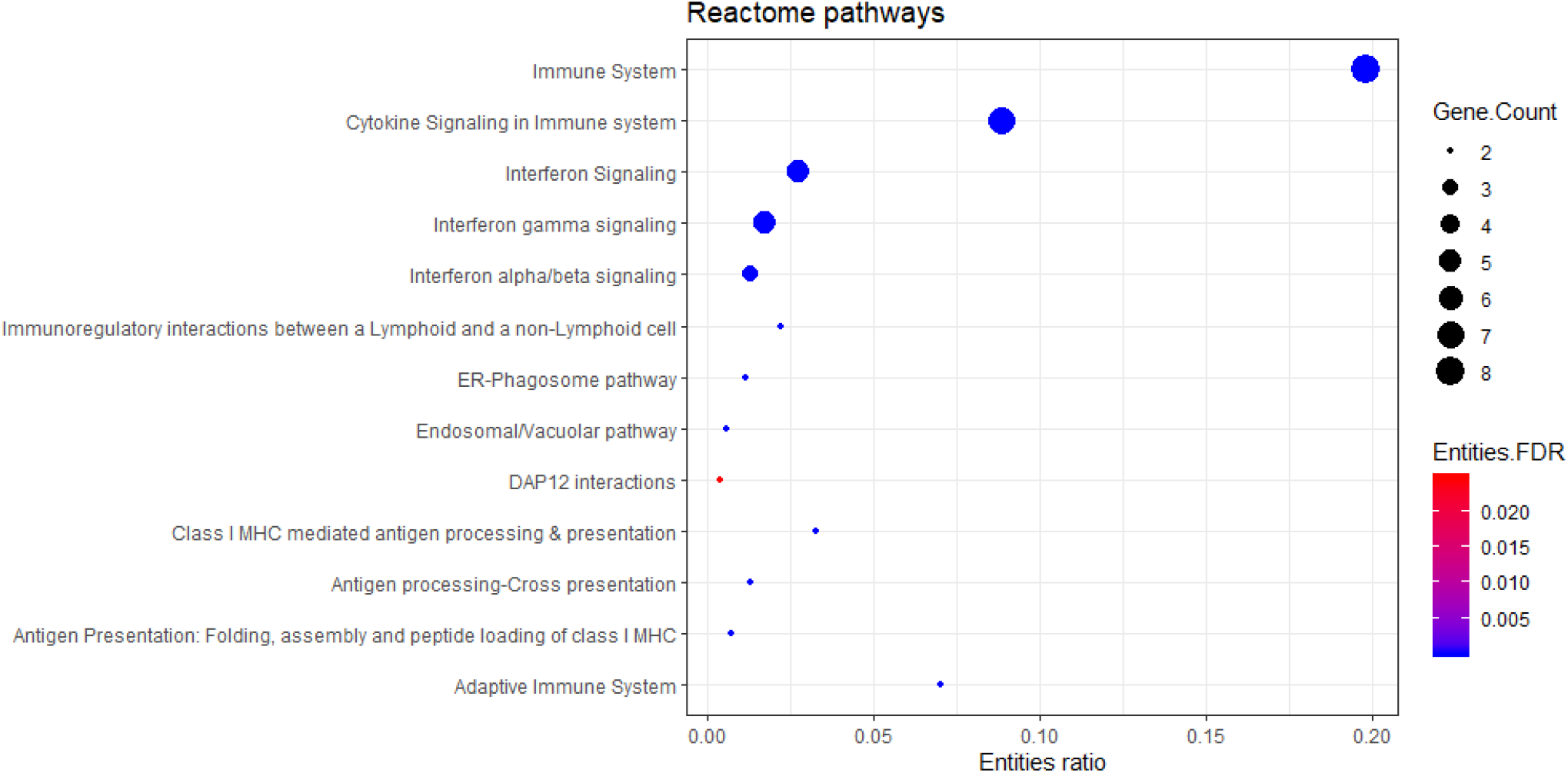
Reactome pathway enrichment analysis of 13 oppositely responding genes (see Figure 4F).

## References

1. Singh P, Arora A, Strand TA, et al. Global Prevalence of Celiac Disease: Systematic Review and Meta-analysis. Clin Gastroenterol Hepatol 2018;16:823-836.e2.

2. Mustalahti K, Catassi C, Reunanen A, et al. The prevalence of celiac disease in Europe: results of a centralized, international mass screening project. Ann Med 2010;42:587–95.

3. Lohi S, Mustalahti K, Kaukinen K, et al. Increasing prevalence of coeliac disease over time. Aliment Pharmacol Ther 2007;26:1217–25.

4. Green PHR, Cellier C. Celiac Disease. N Engl J Med 2007;357:1731–1743.

5. Schuppan D, Junker Y, Barisani D. Celiac disease: from pathogenesis to novel therapies. Gastroenterology 2009;137:1912–33.

6. Sollid LM, Jabri B. Triggers and drivers of autoimmunity: lessons from coeliac disease. Nat Rev Immunol 2013;13:294–302.

7. Valitutti F, Cucchiara S, Fasano A. Celiac Disease and the Microbiome. Nutrients 2019;11.

8. Trynka G, Hunt KA, Bockett NA, et al. Dense genotyping identifies and localizes multiple common and rare variant association signals in celiac disease. Nat Genet 2011;43:1193–201.

9. Jabri B, Sollid LM. T Cells in Celiac Disease. J Immunol 2017;198:3005–3014.

10. Sollid LM, Tye-Din JA, Qiao S-W, et al. Update 2020: nomenclature and listing of celiac disease-relevant gluten epitopes recognized by CD4+ T cells. Immunogenetics 2020;72:85–88.

11. Mäki M. Autoantibodies as markers of autoimmunity in coeliac disease pathogenesis. In: Feighery C, O’Farrelly c, eds. Gastrointestinal immunology and gluten-sensitive disease. Oak Tree Press, Dublin 1994:246–52.

12. Mäki M. The humoral immune system in coeliac disease. Baillieres Clin Gastroenterol 1995;9:231–49.

13. Garber ME, Saldanha A, Parker JS, et al. A B-Cell Gene Signature Correlates With the Extent of Gluten-Induced Intestinal Injury in Celiac Disease. Cell Mol Gastroenterol Hepatol 2017;4:1–17.

14. Pré MF du, Blazevski J, Dewan AE, et al. B cell tolerance and antibody production to the celiac disease autoantigen transglutaminase 2. J Exp Med 2020;217.

15. Garrote JA, Gómez E, León AJ, et al. Cytokine, Chemokine and Immune Activation Pathway Profiles in Celiac Disease: An Immune System Activity Screening by Expression Macroarrays. Drug Target Insights 2008;3:1–11.

16. Bragde H, Jansson U, Fredrikson M, et al. Celiac disease biomarkers identified by transcriptome analysis of small intestinal biopsies. Cell Mol Life Sci 2018;75:4385–4401.

17. Leonard MM, Bai Y, Serena G, et al. RNA sequencing of intestinal mucosa reveals novel pathways functionally linked to celiac disease pathogenesis Sestak K, ed. PLoS One 2019;14:e0215132.

18. Loberman-Nachum N, Sosnovski K, Segni A Di, et al. Defining the Celiac Disease Transcriptome using Clinical Pathology Specimens Reveals Biologic Pathways and Supports Diagnosis. Sci Rep 2019;9:16163.

19. Tutturen AE V, Dørum S, Clancy T, et al. Characterization of the Small Intestinal Lesion in Celiac Disease by Label-Free Quantitative Mass Spectrometry. Am J Pathol 2018;188:1563–1579.

20. Fernandez-Jimenez N, Garcia-Etxebarria K, Plaza-Izurieta L, et al. The methylome of the celiac intestinal epithelium harbours genotype-independent alterations in the HLA region. Sci Rep 2019;9:1298.

21. Lähdeaho ML, Mäki M, Laurila K, et al. Small-bowel mucosal changes and antibody responses after low-and moderate-dose gluten challenge in celiac disease. BMC Gastroenterol 2011;11.

22. Lähdeaho M-L, Kaukinen K, Laurila K, et al. Glutenase ALV003 attenuates gluten-induced mucosal injury in patients with celiac disease. Gastroenterology 2014;146:1649–58.

23. Lähdeaho M-L, Scheinin M, Vuotikka P, et al. Safety and efficacy of AMG 714 in adults with coeliac disease exposed to gluten challenge: a phase 2a, randomised, double-blind, placebo-controlled study. lancet Gastroenterol Hepatol 2019;4:948–959.

24. Taavela J, Viiri K, Popp A, et al. Histological, immunohistochemical and mRNA gene expression responses in coeliac disease patients challenged with gluten using PAXgene fixed paraffin-embedded duodenal biopsies. BMC Gastroenterol 2019;19:189.

25. Comino I, Real A, Vivas S, et al. Monitoring of gluten-free diet compliance in celiac patients by assessment of gliadin 33-mer equivalent epitopes in feces. Am J Clin Nutr 2012;95:670–677.

26. Comino I, Fernández-Bañares F, Esteve M, et al. Fecal Gluten Peptides Reveal Limitations of Serological Tests and Food Questionnaires for Monitoring Gluten-Free Diet in Celiac Disease Patients. Am J Gastroenterol 2016;111:1456–1465.

27. Syage JA, Kelly CP, Dickason MA, et al. Determination of gluten consumption in celiac disease patients on a gluten-free diet. Am J Clin Nutr 2018;107:201–207.

28. Taavela J, Koskinen O, Huhtala H, et al. Validation of morphometric analyses of small-intestinal biopsy readouts in celiac disease. PLoS One 2013;8:e76163.

29. Piñeiro G, Perelman S, Guerschman JP, et al. How to evaluate models: Observed vs. predicted or predicted vs. observed? Ecol Modell 2008;216:316–322.

30. Abadie V, Kim SM, Lejeune T, et al. IL-15, gluten and HLA-DQ8 drive tissue destruction in coeliac disease. Nature 2020;578:600–604.

31. Goodpaster AM, Kennedy MA. Quantification and statistical significance analysis of group separation in NMR-based metabonomics studies. Chemom Intell Lab Syst 2011;109:162–170.

32. Rondanelli, Faliva Gasparri, et al. Micronutrients Dietary Supplementation Advices for Celiac Patients on Long-Term Gluten-Free Diet with Good Compliance: A Review. Medicina (B Aires) 2019;55:337.

33. Juuti-Uusitalo K, Mäki M, Kainulainen H, et al. Gluten affects epithelial differentiation-associated genes in small intestinal mucosa of coeliac patients. Clin Exp Immunol 2007;150:294–305.

34. Senger S, Sapone A, Fiorentino MR, et al. Celiac Disease Histopathology Recapitulates Hedgehog Downregulation, Consistent with Wound Healing Processes Activation Singh SR, ed. PLoS One 2015;10:e0144634.

35. Oittinen M, Popp A, Kurppa K, et al. Polycomb Repressive Complex 2 Enacts Wnt Signaling in Intestinal Homeostasis and Contributes to the Instigation of Stemness in Diseases Entailing Epithelial Hyperplasia or Neoplasia. Stem Cells 2017;35:445–457.

36. Piscaglia AC, Rutella S, Laterza L, et al. Circulating hematopoietic stem cells and putative intestinal stem cells in coeliac disease. J Transl Med 2015;13:220.

37. Freire R, Ingano L, Serena G, et al. Human gut derived-organoids provide model to study gluten response and effects of microbiota-derived molecules in celiac disease. Sci Rep 2019;9:7029.

38. Haber AL, Biton M, Rogel N, et al. A single-cell survey of the small intestinal epithelium. Nature 2017;551:333–339.

39. Nanayakkara M, Lania G, Maglio M, et al. Enterocyte Proliferation and Signaling Are Constitutively Altered in Celiac Disease. PLoS One 2013;8:76006.

40. Ménard S, Cerf-Bensussan N, Heyman M. Multiple facets of intestinal permeability and epithelial handling of dietary antigens. Mucosal Immunol 2010;3:247–259.

41. Matysiak-Budnik T, Moura IC, Arcos-Fajardo M, et al. Secretory IgA mediates retrotranscytosis of intact gliadin peptides via the transferrin receptor in celiac disease. J Exp Med 2008;205:143–54.

42. Kreutz JM, Adriaanse MPM, Ploeg EMC van der, et al. Narrative Review: Nutrient Deficiencies in Adults and Children with Treated and Untreated Celiac Disease. Nutrients 2020;12:500.

43. Vici G, Belli L, Biondi M, et al. Gluten free diet and nutrient deficiencies: A review. Clin Nutr 2016;35:1236–1241.

44. Allen B, Orfila C. The Availability and Nutritional Adequacy of Gluten-Free Bread and Pasta. Nutrients 2018;10:1370.

45. Lerner BA, Phan Vo LT, Yates S, et al. Detection of Gluten in Gluten-Free Labeled Restaurant Food. Am J Gastroenterol 2019;114:792–797.

46. Silvester JA, Comino I, Kelly CP, et al. Most Patients With Celiac Disease on Gluten-free Diets Consume Measurable Amounts of Gluten. Gastroenterology 2019.

47. Silvester JA, Kurada S, Szwajcer A, et al. Tests for Serum Transglutaminase and Endomysial Antibodies Do Not Detect Most Patients With Celiac Disease and Persistent Villous Atrophy on Gluten-free Diets: a Meta-analysis. Gastroenterology 2017;153:689-701.e1.

48. Daveson AJM, Popp A, Taavela J, et al. Baseline quantitative histology in therapeutics trials reveals villus atrophy in most patients with coeliac disease who appear well controlled on gluten-free diet. GastroHep 2020:ygh2.380.

49. Adelman DC, Murray J, Wu T-T, et al. Measuring Change In Small Intestinal Histology In Patients With Celiac Disease. Am J Gastroenterol 2018;113:339–347.

50. Popp A, Mäki M. Gluten-Induced Extra-Intestinal Manifestations in Potential Celiac Disease—Celiac Trait. Nutrients 2019;11:320.

51. Kurppa K, Collin P, Viljamaa M, et al. Diagnosing Mild Enteropathy Celiac Disease: A Randomized, Controlled Clinical Study. Gastroenterology 2009;136:816–823.

52. Leo L De, Bramuzzo M, Ziberna F, et al. Diagnostic accuracy and applicability of intestinal auto-antibodies in the wide clinical spectrum of coeliac disease. EBioMedicine 2020;51:102567.

53. Choung RS, Khaleghi S, Cartee AK, et al. Community-Based Study of Celiac Disease Autoimmunity Progression in Adults. Gastroenterology 2020;158:151-159.e3.

54. Yin X, Farin HF, Es JH van, et al. Niche-independent high-purity cultures of Lgr5+ intestinal stem cells and their progeny. Nat Methods 2014;11:106–112.

55. Uspenskaya ID, Shirokova NY. Duodenal mucosa in children with coeliac disease in catamnesis and varying compliance with the gluten-free diet. Bratislava Med J 2014;115:150–155.

56. Ciacci C, Vizio D Di, Seth R, et al. Selective reduction of intestinal trefoil factor in untreated coeliac disease. 2002.

57. Pietz G, De R, Hedberg M, et al. Immunopathology of childhood celiac disease-Key role of intestinal epithelial cells. PLoS One 2017;12:e0185025.

58. Forsberg G, Fahlgren A, Hörstedt P, et al. Presence of bacteria and innate immunity of intestinal epithelium in childhood celiac disease. Am J Gastroenterol 2004;99:894–904.

59. Capaldo CT, Beeman N, Hilgarth RS, et al. IFN-γ and TNF-α-induced GBP-1 inhibits epithelial cell proliferation through suppression of β-catenin/TCF signaling. Mucosal Immunol 2012;5:681–90.

60. Kim YS, Ho SB. Intestinal goblet cells and mucins in health and disease: recent insights and progress. Curr Gastroenterol Rep 2010;12:319–30.

61. Wang ML, Keilbaugh SA, Cash-Mason T, et al. Immune-mediated signaling in intestinal goblet cells via PI3-kinase-and AKT-dependent pathways. Am J Physiol - Gastrointest Liver Physiol 2008;295.

62. Lahdenperä A, Ludvigsson J, Fälth-Magnusson K, et al. The effect of gluten-free diet on Th1-Th2-Th3-associated intestinal immune responses in celiac disease. Scand J Gastroenterol 2011;46:538–549.

63. Corazza GR, Villanacci V, Zambelli C, et al. Comparison of the Interobserver Reproducibility With Different Histologic Criteria Used in Celiac Disease. Clin Gastroenterol Hepatol 2007;5:838–843.

64. Mubarak A, Nikkels P, Houwen R, et al. Reproducibility of the histological diagnosis of celiac disease. Scand J Gastroenterol 2011;46:1065–1073.

65. Arguelles-Grande C, Tennyson CA, Lewis SK, et al. Variability in small bowel histopathology reporting between different pathology practice settings: impact on the diagnosis of coeliac disease. J Clin Pathol 2012;65:242–247.

66. Picarelli A, Borghini R, Donato G, et al. Weaknesses of histological analysis in celiac disease diagnosis: new possible scenarios. Scand J Gastroenterol 2014;49:1318–1324.

67. Werkstetter KJ, Korponay-Szabó IR, Popp A, et al. Accuracy in Diagnosis of Celiac Disease Without Biopsies in Clinical Practice. Gastroenterology 2017;153:924–935.

68. Charlesworth RPG, Agnew LL, Scott DR, et al. Celiac disease gene expression data can be used to classify biopsies along the Marsh score severity scale. J Gastroenterol Hepatol 2019;34:169–177.

